# The Burden of Cancer and Pre-cancerous Conditions Among Transgender Individuals in a Large Healthcare Network

**DOI:** 10.1101/2024.03.24.24304777

**Authors:** Shuang Yang, Yongqiu Li, Christopher W. Wheldon, Mattia Prosperi, Thomas J. George, Elizabeth A. Shenkman, Fei Wang, Jiang Bian, Yi Guo

## Abstract

The current study aimed to examine the prevalence of and risk factors for cancer and pre-cancerous conditions, comparing transgender and cisgender individuals, using 2012-2023 electronic health record data from a large healthcare system. We identified 2,745 transgender individuals using a previously validated computable phenotype and 54,900 matched cisgender individuals. We calculated the prevalence of cancer and pre-cancer related to human papillomavirus (HPV), human immunodeficiency virus (HIV), tobacco, alcohol, lung, breast, colorectum, and built multivariable logistic models to examine the association between gender identity and the presence of cancer or pre-cancer. Results indicated similar odds of developing cancer across gender identities, but transgender individuals exhibited significantly higher risks for pre-cancerous conditions, including alcohol-related, breast, and colorectal pre-cancers compared to cisgender women, and HPV-related, tobacco-related, alcohol-related, and colorectal pre-cancers compared to cisgender men. These findings underscore the need for tailored interventions and policies addressing cancer health disparities affecting the transgender population.

## Introduction

Transgender persons are people whose gender identity or expression does not align with society’s expectations based on the sex recorded at birth^1^. Global estimates suggest that 0.3% to 0.5% of adults self-identify as transgender^2^. A recent survey has reported that as high as 1.6% of the general U.S. adults identify as transgender^3^. Transgender individuals consistently experience disproportionately high prevalence of adverse health outcomes, such as the human immunodeficiency virus (HIV), sexually transmitted infections, mental health distress, and substance abuse^4^. Despite the growing literature on the unique health challenges faced by transgender individuals, our understanding of cancer burden and cancer risk in this vulnerable population remains sparse^5^, primarily due to the unavailability of relevant data^6^. Reluctance to self-identify and participate in health surveys, coupled with the scarcity of inclusion of gender identity measures in national surveillance efforts, pose challenges in obtaining population-based representative samples and conducting longitudinal studies to examine cancer burden in this population. Current insights into cancer risks in transgender populations rely mainly on anecdotal evidence and small-scale studies^7,8^, resulting in inconsistent findings.

Evidence suggests transgender persons may face elevated risk of certain cancers compared to the general population, including breast cancer^7^, cancers linked to the human papillomavirus (HPV)^9^ and HIV^6^, and those related to tobacco and alcohol use^10,11^. First, some transgender individuals choose to undergo the regimen of hormone therapy, which includes prolonged use of high doses of exogenous estrogens and androgen blockers for breast development histologically similar to those of biological females^12^, potentially increasing the risk of breast cancer^13,14^. Additionally, nationwide surveys in the U.S. have shown that transgender individuals are twice as likely to smoke cigarettes compared to cisgender individuals^15^. Concurrently, a recent systematic review has revealed a high prevalence of hazardous alcohol use in the transgender population^16^. These behaviors amplify the risk for cancers associated with smoking and drinking^17^, given that both are well-documented, modifiable risk factors for cancer^15,16^. Moreover, transgender individuals’ greater engagement in risky sexual behaviors^18–21^, such as unprotected sex, oral and anal intercourse, and participation in sex work, escalates their vulnerability to infections by HPV and HIV. These infections further increase the risk for cancers associated with these viruses. An observational study found that transgender individuals assigned female at birth had a higher cervicovaginal HPV prevalence compared to the cisgender women (30.5% vs. 20.0%)^22^. In addition, the prevalence of HIV is reported to be disproportionately high at 13.7% among transgender individuals, including a higher rate of 18.8% among transgender women and 2.0% among transgender men^21^.

Pre-cancer, also known as pre-cancerous lesions or conditions, refers to abnormal cellular and tissue changes that have the potential to develop into cancer if untreated^23^. A systematic review and meta-analysis of studies regarding oral pre-cancerous lesions revealed that the rate of malignant transformation varies widely, from less than 1% to over 50%, depending on the type and location of the lesion^24^. Various types of pre-cancerous conditions exist, each posing a specific risk of progressing to cancer in particular organs. For instance, cervical intraepithelial neoplasia (CIN), a well-known HPV-related pre-cancerous lesion, can progress to invasive cervical cancer without intervention^25^. Investigating the burden pre-cancer in transgender individuals is crucial for several reasons. First, many transgender individuals are young, and although cancer prevalence in this group may be low, a significant proportion may harbor pre-cancerous lesions, indicating a heightened future cancer risk. Second, transgender individuals often encounter unique challenges and barriers to healthcare access, potentially leading to delays in pre-cancer diagnosis and treatment. Thus, studying pre-cancer prevalence and risk factors in transgender individuals can inform interventions to address cancer disparities and enhance health outcomes in this population.

Electronic health record (EHR) data has emerged as a crucial tool for addressing knowledge gaps and overcoming challenges in cancer research within the transgender community. The widespread adoption of EHRs, along with the establishment of clinical research networks, has made large-scale, longitudinal clinical data available. Given their rich clinical data including structured information such as diagnoses and procedures, as well as unstructured clinical narratives, EHRs offer a valuable resource for identifying transgender populations and the cancer and pre-cancer cases among them. Moreover, EHRs provide extensive population coverage, enabling the identification of sizable cisgender individuals (i.e., controls), thus allow comparative analysis that reveals cancer disparities. Additionally, compared to cancer registries, EHRs comprise detailed and comprehensive list of cancer risk factors. These factors are crucial for understanding unique cancer burdens among transgender individuals. In this study, we aimed to examine the prevalence of and risk factors for cancer and pre-cancer conditions, comparing transgender and cisgender individuals using EHRs data. Our hypothesis is that transgender individuals are at a heightened risk for cancer and pre-cancer compared to cisgender individuals.

## Methods

### Data Source and Study Population

This study was approved by the University of Florida (UF) Institutional Review Board. We obtained 2012-2023 patient-level EHR data from the UF Health Integrated Data Repository (IDR), a clinical data warehouse that aggregates data from UF’s various clinical and administrative information systems, including the Epic EHR system. The IDR contains more than 1 billion observational facts pertaining to more than 2 million patients. Our study population included a cohort of transgender individuals and a matching cohort of cisgender individuals identified in the UF Health EHRs. The transgender individuals were identified using a computable phenotype algorithm which we had developed and validated previously using UF Health EHRs^26^. The algorithm classified individuals as transgender if they met the following criteria: 1) recorded gender identity as transgender; or 2) had at least one transgender diagnosis code from the diagnosis table, along with at least one transgender keyword from clinical notes. The CP algorithm achieved a 0.955 F1-score on the training data and a perfect F1-score on the independent testing data.

The index date for transgender cohort was defined as the date of the first recorded evidence of transgender status. To create a cisgender cohort, each transgender individual was matched with 10 cisgender men and 10 cisgender women randomly selected from the UF Health EHRs, based on age and calendar year of the index date. For the cisgender cohort, the index date was defined as a randomly selected diagnosis date. We excluded individuals who: 1) had an age at first encounter of less than three years; 2) had no encounter after the index date; and 3) had a gap of longer than two years between any two encounters.

### Study Outcomes

The primary study outcomes were incident cases of HPV-related, HIV-related, tobacco-related, and alcohol-related cancers among the study population^27–30^. We also examined the incident cases of lung, breast, and colorectal cancer separately because clear screening guidelines for primary prevention have been published for these cancers. HPV-related cancers included oropharyngeal, cervical, anal, vaginal, vulvar, and penile cancers^27^. HIV-related cancers included Kaposi sarcoma, non-Hodgkin lymphoma, and cervical cancer^28^. Tobacco-related cancers included lung, head and neck, liver, esophagus, bladder, kidney, stomach, pancreas, colorectal, and cervical cancers^30^. Alcohol-related cancers included head and neck, esophageal, liver, breast, and colorectal cancers^29^. Additionally, the incident cases of any cancer, covering all 18 individual cancer sites mentioned above, were calculated. We identified patients with these cancers in UF Health EHRs using International Classification of Diseases (ICD)-9/10 CM codes.

The secondary study outcomes were incident cases of pre-cancer diseases for the 18 cancers among the study population. We defined the pre-cancers lesions, conditions, and diseases based on criteria established by the Canadian Cancer Society^23^ and related literature reviews^31–42^. Similar to the cancer cases, we grouped the pre-cancer diseases into pre-HPV-related, pre-tobacco-related, pre-alcohol-related, pre-lung, pre-breast, and pre-colorectal cancers (Supplemental Table 1).

**Table 1.** Distributions of demographic characteristics by gender identity.

| Variables | Transgender<br>(N=2,745) | Women<sup>a</sup><br>(N=27,450) | P -<br>value | Men<sup>a</sup><br>(N=27,450) | P -<br>value |
| --- | --- | --- | --- | --- | --- |
| Age at index date | 25.1 (14.0) | 25.1 (14.0) | 1.00 | 25.1 (14.0) | 1.00 |
| Race-Ethnicity<sup>b</sup> |  |  |  |  |  |
| NHW | 1,803 (65.7) | 14,675 (53.5) | < 0.001 | 15,307 (55.8) | < 0.001 |
| NHB | 284 (10.4) | 6,344 (23.1) |  | 5,859 (21.3) |  |
| NHO | 181 (6.6) | 1,925 (7.0) |  | 1,882 (6.9) |  |
| Hispanics | 242 (8.8) | 3,119 (11.4) |  | 2,699 (9.8) |  |
| Unknown | 235 (8.6) | 1,387 (5.1) |  | 1,703 (6.2) |  |
| Insurance payer |  |  |  |  |  |
| Private | 1,147 (41.8) | 13,948 (50.8) | < 0.001 | 13,837 (50.4) | < 0.001 |
| Medicaid and others<sup>c</sup> | 539 (19.6) | 9,708 (35.4) |  | 8,772 (32.0) |  |
| Medicare | 181 (6.6) | 1,261 (4.6) |  | 1,351 (4.9) |  |
| Self-pay | 132 (4.8) | 1,516 (5.5) |  | 2,292 (8.4) |  |
| Unknown | 746 (27.2) | 1,017 (3.7) |  | 1,198 (4.4) |  |
| Smoking status |  |  |  |  |  |
| Current smoker | 289 (10.5) | 1,792 (6.5) | < 0.001 | 2,932 (10.7) | < 0.001 |
| Former smoker | 302 (11.0) | 2,196 (8.0) |  | 2,409 (8.8) |  |
| Never smoker | 1,569 (57.2) | 19,656 (71.6) |  | 17,330 (63.1) |  |
| Unknown | 585 (21.3) | 3,806 (13.9) |  | 4,779 (17.4) |  |
| Family cancer history |  |  |  |  |  |
| No | 2,726 (99.3) | 27,034 (98.5) | < 0.001 | 27,245 (99.2) | 0.750 |
| Yes | 19 (0.7) | 416 (1.5) |  | 205 (0.8) |  |
| CCI<sup>d</sup> |  |  |  |  |  |
| 0 | 2,154 (78.5) | 22,321 (81.3) | < 0.001 | 21,935 (79.9) | < 0.001 |
| 1 | 400 (14.6) | 3,981 (14.5) |  | 4,234 (15.5) |  |
| ≥ 2 | 191 (7.0) | 1,148 (4.2) |  | 1,281 (4.7) |  |
| Number of Outpatient visits (Mean, SD)<sup>e</sup> | 3.2, 6.1 | 4.3, 6.8 | < 0.001 | 3.5, 6.9 | 0.048 |
| Number of Inpatient visits (Mean, SD)<sup>e</sup> | 0.15, 0.6 | 0.15, 0.5 | 0.500 | 0.2, 0.6 | < 0.001 |

<sup>a</sup>1:10 matched to the transgender individuals. <sup>b</sup>NHW: Non-Hispanic White; NHB: Non-Hispanic Black; NHO: Non-Hispanic Other. <sup>c</sup>Other insurance: charity and worker's compensation. <sup>d</sup>CCI: Charlson comorbidity index; 0 = no comorbidity, 1 = some comorbidities, ≥ 2 = a substantial burden of comorbidities. <sup>e</sup>SD: standard deviation. Means and SDs were calculated using data within 12 months prior to the index date.

### Covariates

Covariates included age at index date, race-ethnicity, insurance payer, smoking status, family history of cancer, health care utilization (i.e., outpatient and inpatient visits), and Charlson comorbidity index (CCI). Race-ethnicity was categorized as Non-Hispanic White (NHW), Non-Hispanic Black (NHB), Non-Hispanic Other (NHO), Hispanics, or Unknown. Insurance payer was categorized as Medicare, private insurance, self-pay, and Medicaid or other insurance (e.g., charity, worker’s compensation). Smoking status was determined using the most recent EHRs before the index date and categorized as current, former, never smoker, or unknown. Additionally, baseline measures of healthcare utilization, family cancer history (ICD-9: V16; ICD-10: Z80) and CCI were extracted from EHR data within 12 months prior to the index date. Healthcare utilization was measured by the numbers of outpatient and inpatient visits. Family history of cancer and conditions in the CCI (e.g., diabetes) were identified using relevant ICD-9/10-CM codes and confirmed with at least one inpatient or outpatient diagnosis. We calculated the CCI following the modified algorithm by Klabunde et al.^43^.

### Statistical Analysis

First, we calculated summary statistics to characterize the individuals in our study population by gender identity. Next, we calculated and compared the prevalence rates of cancer and pre-cancer cases in the transgender vs. cisgender male and female cohorts. Prevalence rate was defined as the number of incident cases of cancer or pre-cancer divided by the number of total individuals in a study cohort^44^. Differences in the distributions of patient characteristics and cancer and pre-cancer cases between transgender vs. cisgender male and female cohorts were tested using t-tests for continuous variables and Chi-squared or Fisher’s exact tests for categorical variables. Lastly, adjusting for the covariates, we built multivariable logistic regression models to compare the odds of developing each type of cancer and pre-cancer across the transgender and cisgender cohorts. We also examined whether cancer or pre-cancer risk differed by gender identify by including and testing interactions between gender identify and the covariates in the logistic models. All effects were estimated by odds ratios (ORs) with 95% confidence intervals (CIs). Two-sided p-values were calculated for all statistics considering a significance level of 0.05. Data processing and management were conducted using python 3.9.4. Statistical analyses were conducted using SAS 9.4 (SAS Institute Inc., Cary, NC, USA).

## Results

We identified 2,745 transgender individuals with 27,450 matched cisgender women and 27,450 match cisgender men in the UF Health EHRs. Table 1 displays the distributions of the patient characteristics stratified by gender identity. After matching, the mean age was the same (25.1 years) across the transgender and cisgender female and male cohorts (*p* = 1.000). Beyond age, we observed a significant difference in each of the characteristic by gender identity. Compared to the cisgender individuals, the transgender individuals were more likely to be NHW (65.7% in transgender vs. 53.5% and 55.8% in cisgender women and men, *p* < 0.001) and have a substantial burden of comorbidities (7.0% in transgender vs. 4.2% and 4.7% in cisgender women and men, *p* < 0.001). In contrast, the transgender individuals were less likely to have private insurance (41.8% in transgender vs. 50.8% and 50.4% in cisgender women and men, *p* < 0.001), be a never smoker (57.2% in transgender vs. 71.6% and 63.1% in cisgender women and men, *p* < 0.001), and have outpatient visits (3.2 times in transgender vs. 4.3 and 3.5 times in cisgender women and men in the past year, *p* < 0.001). Further, the transgender individuals were less likely to have family cancer history than cisgender women (0.7% vs. 1.5%, *p* < 0.001) and had fewer number of inpatient visits in the past year than cisgender men (0.15 vs. 0.2, *p* < 0.001).

We summarized the prevalence rates of cancer and pre-cancer cases in our study populations stratified by gender identity in Table 2. As shown in the table, the prevalence of any cancer in the transgender individuals was comparable to that in the cisgender women (1.0% vs. 1.3%, *p* = 0.128) and men (1.0% vs. 1.1%; *p* = 0.636). Overall, the prevalence of cancer was not statistically different between the transgender and cisgender individuals, except that the transgender individuals had a significantly higher rate of breast cancer than the cisgender men (0.3% vs. 0.004%, *p* < 0.001). Regarding the pre-cancer cases, the prevalence of any pre-cancer was higher in the transgender individuals than the cisgender men (15.0% vs. 12.4%; *p* < 0.001) and women (15.0% vs. 13.9%; *p* = 0.134), although the latter comparison was statistically non-significant. Compared to the cisgender men, the transgender individuals had a significantly higher prevalence of HPV-related pre-cancers (3.4% vs. 2.1%, *p* < 0.001), alcohol-related pre-cancers (7.5% vs. 5.5%, *p* < 0.001), pre-breast cancer (1.0% vs. 0.01%, *p* < 0.001), and pre-colorectal cancer (6.7% vs. 5.4%, *p* < 0.001). Compared to the cisgender women, the transgender individuals had a significantly higher prevalence of alcohol-related pre-cancers (7.5% vs. 6.5%, *p* = 0.048) and pre-breast cancer (1.5% vs. 1.0%, *p* = 0.032).

**Table 2.** Prevalence rates of cancer and pre-cancer cases by gender identity.

| Conditions | Transgender<br>(N=2,745) | Women<br>(N=27,450) | *p*-value | Men<br>(N=27,450) | *p*-value |
| --- | --- | --- | --- | --- | --- |
| **Cancer** |  |  |  |  |  |
| Any cancer<sup>a</sup> | 28 (1.0%) | 358 (1.3%) | 0.128 | 314 (1.1%) | 0.636 |
| HPV-related<sup>b</sup> | 10 (0.4%) | 69 (0.3%) | 0.242 | 69 (0.3%) | 0.242 |
| HIV-related<sup>c</sup> | 8 (0.3%) | 92 (0.3%) | 0.704 | 87 (0.3%) | 1.000 |
| Tobacco-related<sup>d</sup> | 15 (0.6%) | 168 (0.6%) | 0.796 | 219 (0.8%) | 0.170 |
| Alcohol-related<sup>e</sup> | 12 (0.4%) | 185 (0.7%) | 0.170 | 116 (0.4%) | 0.877 |
| Lung | 9 (0.3%) | 53 (0.2%) | 0.178 | 81 (0.3%) | 0.713 |
| Breast | 7 (0.3%) | 145 (0.5%) | 0.064 | 1 (0.004%) | < 0.001 |
| Colorectal | 4 (0.2%) | 45 (0.2%) | 1.000 | 41 (0.2%) | 1.000 |
| **Pre-cancer** |  |  |  |  |  |
| Any pre-cancer<sup>a</sup> | 411 (15.0%) | 3,823 (13.9%) | 0.134 | 3,409 (12.4%) | < 0.001 |
| HPV-related<sup>b</sup> | 92 (3.4%) | 961 (3.5%) | 0.743 | 584 (2.1%) | < 0.001 |
| Tobacco-related<sup>d</sup> | 351 (12.8%) | 3,505 (12.8%) | 0.976 | 3,166 (11.5%) | 0.053 |
| Alcohol-related<sup>e</sup> | 206 (7.5%) | 1,784 (6.5%) | 0.048 | 1,519 (5.5%) | < 0.001 |
| Lung | 16 (0.6%) | 155 (0.6%) | 0.894 | 109 (0.4%) | 0.158 |
| Breast | 41 (1.5%) | 286 (1.0%) | 0.032 | 2 (0.01%) | < 0.001 |
| Colorectal | 184 (6.7%) | 1,728 (6.3%) | 0.411 | 1,472 (5.4%) | 0.004 |

<sup>a</sup>Any of the 18 cancer types: Kaposi sarcoma, non-Hodgkin lymphoma, cervical cancer, oropharyngeal, anal, vaginal, vulvar, penile, lung, head and neck, liver, esophagus, bladder, kidney, stomach, pancreas, colorectal and breast cancer. <sup>b</sup>HPV-related cancers include oropharyngeal, cervical, anal, vaginal, vulvar, and penile cancers. <sup>c</sup>HIV-cancer include Kaposi sarcoma, non-Hodgkin lymphoma and cervical cancer. <sup>d</sup>Tobacco-related cancers include lung, head and neck, liver, esophagus, bladder, kidney, stomach, pancreas, colorectal, and cervical cancers.
<sup>e</sup>Alcohol-related cancers include head and neck, esophageal, liver, breast, and colorectal cancers.

We summarized the main findings from the multivariable logistic regression models examining the association of gender identity with cancer and pre-cancer cases in Figure 1. For the cancer cases, the odds of developing any cancer or each of the cancer types was statistically the same across the transgender and cisgender cohorts, as none of the ORs for gender identity were statistically significant adjusting for the covariates. However, the transgender individuals were significantly more likely to develop any pre-cancer than the cisgender women (OR = 1.25; 95% CI = 1.11 – 1.43) and men (OR = 1.43; 95% CI = 1.25 – 1.67). Compared to the cisgender women, the transgender individuals were more likely to develop alcohol-related pre-cancers (OR = 1.25; 95% CI = 1.11 – 1.43), pre-breast cancer (OR = 2.0; 95% CI = 1.67 – 3.33), and pre-colorectal cancer (OR = 1.25; 95% CI = 1.11 – 1.43). Compared to the cisgender men, the transgender individuals were more likely to develop HPV-related pre-cancers (OR = 1.67; 95% CI = 1.25 – 2.50), tobacco-related pre-cancers (OR = 1.25; 95% CI = 1.11 – 1.43), alcohol-related pre-cancers (OR = 1.67; 95% CI = 1.25 – 1.67), and pre-colorectal cancer (OR =1.43; 95% CI = 1.25 – 1.67), adjusting for the covariates.

**Figure 1.**
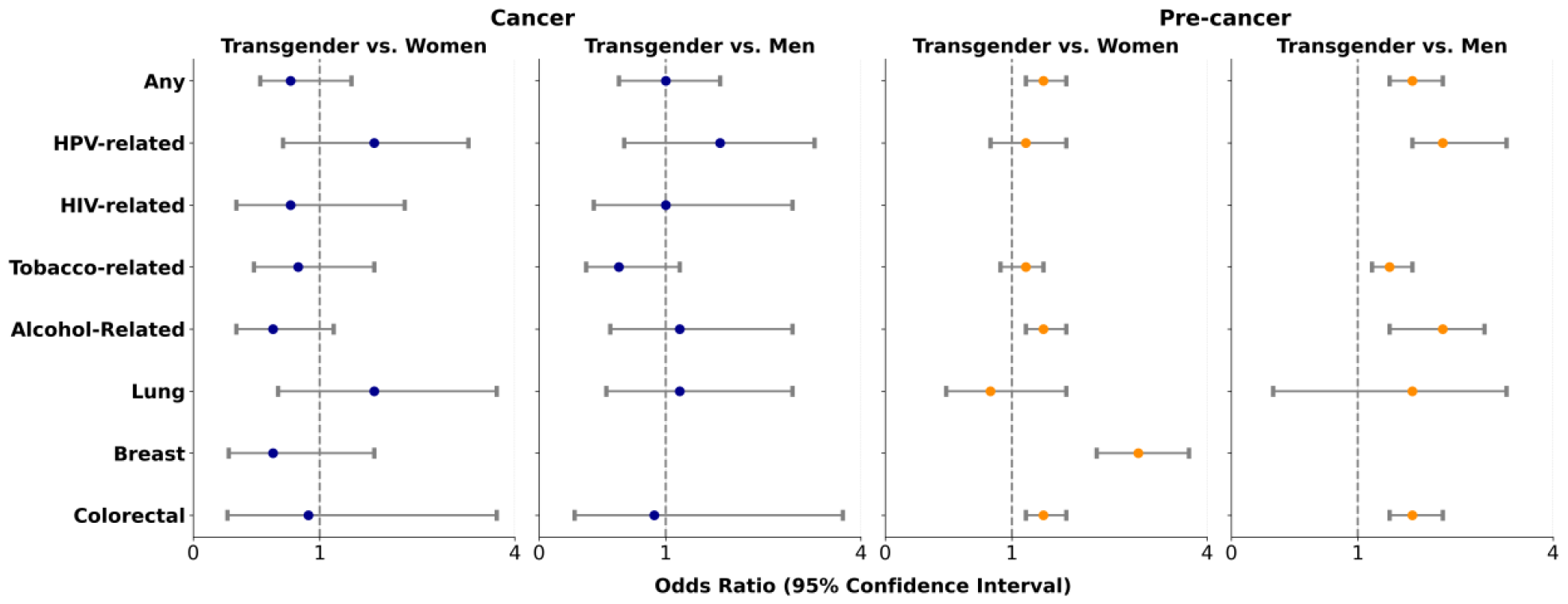
Adjusted odds ratios estimating the association of gender identity with cancer and pre-cancer cases.

As the risk for the pre-cancers was found to differ significantly by gender identity, we further examined whether the effects of the cancer risk factors also differed by gender identity for any pre-cancer as well as HPV-related, alcohol-related, and tobacco-related pre-cancers. We plotted the odds ratios from the multivariable logistic regression models examining the interaction effects between gender identity and the cancer risk factors in Figure 2. Having private insurance or Medicare was a protective factor for pre-cancers, and the protective effect was greater in transgender as compared to cisgender individuals. Being NHB, as compared to being NHW, was a significant risk factor for any pre-cancer in transgender individuals only. For all the pre-cancer typed analyzed, being NHB was a greater risk factor for transgender as compared to cisgender individuals. Being Hispanic was a significant risk factor for the pre-cancers in cisgender individuals only, with the risk being higher in men compared to women. Having a substantial burden of comorbidities (CCI ≥ 2 vs. = 0) was a greater risk factor for transgender individuals for any pre-cancer and tobacco-related pre-cancer. Regarding smoking status, being former smoker as compared to never smoker was a significant risk factor for HPV-related pre-cancers in cisgender individuals, with the risk being much higher in women than men. The effects of outpatient and inpatient visits on any pre-cancer risk were lower in transgender as compared to cisgender individuals.

**Figure 2.**
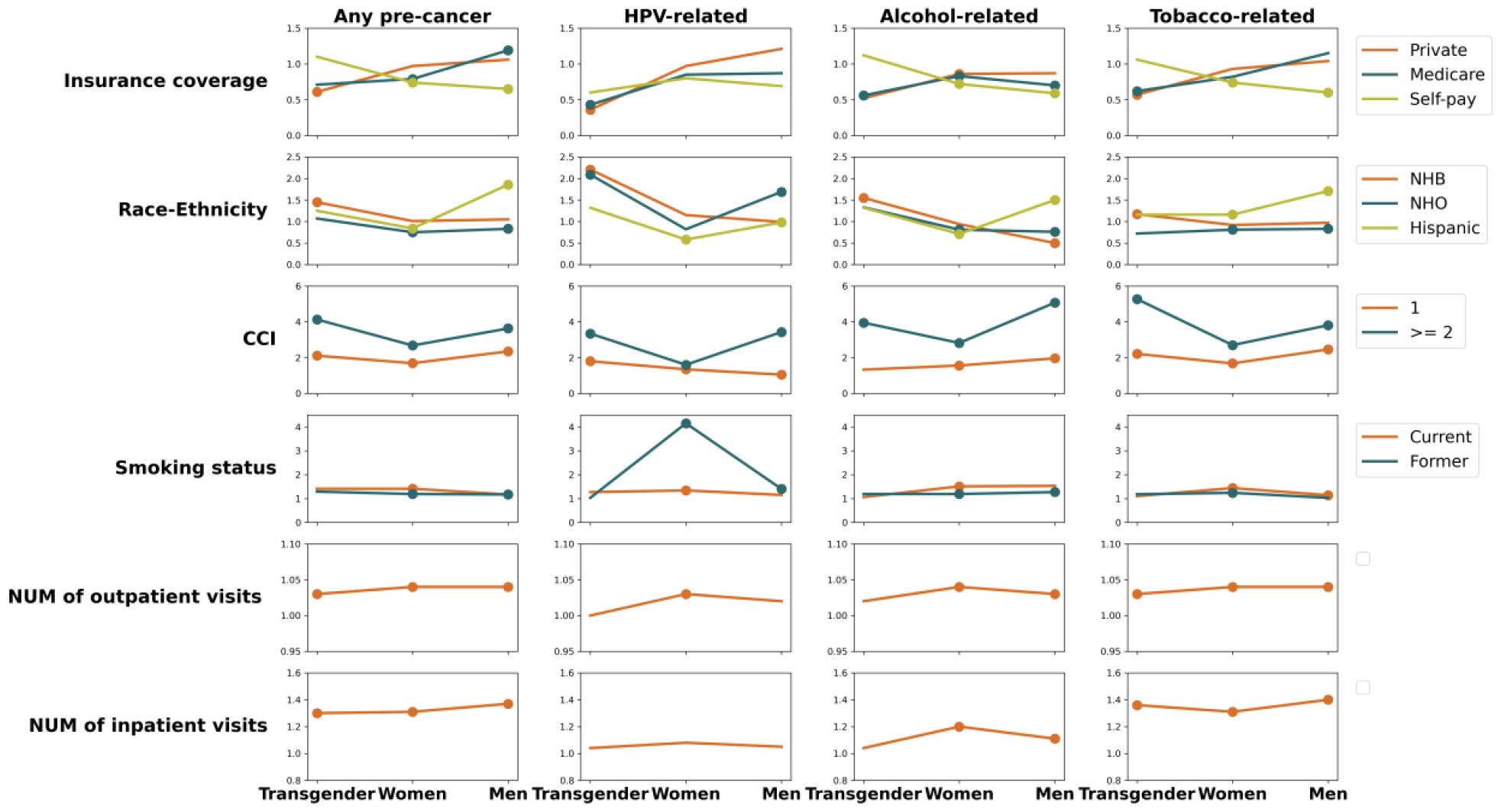
Interaction plots of gender identity with cancer risk factors for pre-cancer diseases. NHB: Non-Hispanic Black; NHO: Non-Hispanic Other. CCI: Charlson comorbidity index; 0 = no comorbidity, 1 = some comorbidities, ≥ 2 = a substantial burden of comorbidities. Dots represent statistically significant odds ratios. Reference groups are Medicaid for insurance coverage, non-Hispanic White for race-ethnicity, 0 for CCI, never smoker for smoking status.

## Discussion and Conclusions

In this study, we utilized EHRs data from a large healthcare system to examine the risk of cancer and pre-cancer and explore the impact of cancer risk factors across different gender identities. Our findings indicate that while the odds of developing any cancer were similar between transgender and cisgender individuals, transgender individuals were more likely to develop the pre-cancer diseases than their cisgender counterparts. Specifically, transgender individuals showed elevated risks for alcohol-related, breast, and colorectal pre-cancers compared to cisgender women, and higher risks for HPV-related, tobacco-related, alcohol-related, and colorectal pre-cancers compared to cisgender men in multivariable analysis. Furthermore, our analysis identified having private insurance or Medicare as protective factors against pre-cancers in transgender individuals, while factors such as being NHB or having a substantial burden of comorbidities emerged as significant risk factors.

Our study revealed that the odds of developing cancer were not significantly different between transgender and cisgender individuals, which does not align with our hypothesis that transgender individuals are at higher risk for cancer. A plausible reason for this non-significant relationship is that the sample sizes of the cancer cases were small (i.e., low prevalence rates of cancer), which did not fully capture the variances in cancer risks. The transgender cohort in our study was relatively young (mean age = 25.1), whereas cancer risk increases with age, with most cases being observed in people aged 50 and over^45^. Additionally, potential underdiagnosis among transgender individuals, possibly resulting from systemic barriers to healthcare access, could lead to an underestimation of cancer cases in this group. On the other hand, we observed an increased risk of developing pre-cancerous conditions in transgender as compared to cisgender individuals, which supports our initial hypothesis. This paradox indicates that although age may protect against cancer, transgender individuals are still at a higher risk for early-stage carcinogenic processes. Such early indicators of cancer risk might be exacerbated by biological factors, such as prolonged use of hormone therapy, and lifestyle factors, such as increased rates of smoking and alcohol consumption. Our findings emphasize the importance of developing health interventions against cancer tailored to the transgender community’s unique needs, focusing on modifiable risk factors such as smoking cessation and alcohol moderation, as well as enhancing access to preventative healthcare services including regular cancer screenings and HPV vaccination programs. By addressing these factors, healthcare providers can work towards reducing the burden of cancer and pre-cancer in transgender individuals, ultimately improving their overall health outcomes and quality of life.

Our analysis also revealed several risk factors associated with pre-cancer risk among transgender individuals, uncovering subtle differences in disparities. For instance, transgender individuals with private insurance or Medicare exhibited lower pre-cancer risks compared to those with Medicaid. Transgender individuals with Medicaid insurance may face increased risk due to limited access to specialized care or preventative service. For example. prior research indicates that Medicaid enrollees had lower rates of breast, cervical, and colorectal cancer screening compared to those with commercial insurance^46^. Additionally, NHB transgender individuals demonstrated elevated pre-cancer risks compared to their NHW counterparts, possibly due to systemic inequalities in healthcare access and utilization, as well as higher rates of underlying health conditions in NHB. According to the National Transgender Discrimination Survey (NTDS), Black transgender adults in the U.S. have higher rate unemployed, living in poverty, have been sexually assaulted, had negative experience with a healthcare provider, living with HIV than cisgender black people^47^. Furthermore, the presence of substantial comorbidities emerged as a significant predictor of pre-cancer risk among transgender individuals, reflecting the complex interplay between health status and cancer susceptibility. For example, conditions such as HIV, often prevalent in transgender populations, can weaken immune function, contributing individuals to pre-cancerous lesions. To tackle these differences, we need specific actions, including improving the availability of specialized care or preventative service in Medicaid coverage for transgender individuals, implementing culturally competent healthcare practices within NHB communities, and addressing underlying health issues to reduce pre-cancer risk.

Our study has several strengthens, including the use of comprehensive EHR data, facilitating a large-scale examination of cancer and pre-cancer conditions among transgender individuals, and the inclusion of matched cisgender cohorts enhances the validity of comparative analysis. Our study also has several limitations. Although our transgender computable phenotype could identify transgender subgroups (i.e., transgender men and women), we did not analyze these subgroups separately due to small sample sizes of cancer cases. Furthermore, we were unable to examine certain cancer risk factors potentially important for the transgender population, such as sexual behaviors and alcohol use, due to the lack of these data in EHRs.

In conclusion, our study provides valuable insights into the cancer and pre-cancer burden in the transgender population. The findings highlight the urgent need for targeted interventions and healthcare policies aimed at reducing cancer health disparities in this vulnerable population. Future research efforts should focus on elucidating the underlying mechanisms driving cancer risks among transgender individuals including the role of hormone therapy, behavioral risk factors, and barriers to accessing quality healthcare.

## Data Availability

Please include a statement regarding the availability of all data referred to in the manuscript.

## Acknowledgement

This study was supported by grants R21CA245858 and R21CA245858-01A1S1 from the National Institutes of Health (NIH). This study was also partially supported by NIH grants R01CA246418, R01CA246418-02S1, R21CA253394-01A1, R01AG080624, and R21AG068717. The authors wish to thank the Cancer Informatics Shared Resource in the UF Health Cancer Center for data analytics support.

---

**Supplemental Table 1.** Cancer sites and corresponding pre-cancer conditions.

| Cancer site | Pre-cancer |
| --- | --- |
| Oropharyngeal<sup>48</sup>, Head and neck<sup>49</sup> cancer | Leukoplakia; Erythroplakia; Oral submucosal fibrosis; Oral lichen planus; Premalignant mucosal changes; Plummer-Vinson; Fanconi anemia and dyskeratosis congenita; Palatal lesions; Actinic keratosis |
| Colorectal<sup>50</sup> | Familial adenomatous polyposis; Tubular adenomas; Villous adenomas; Tubulovillous adenomas; Hyperplastic polyposis; Cowden syndrome; Lynch syndrome; Peutz-Jeghers syndrome; Juvenile polyposis syndrome; Inflammatory Bowel Disease |
| Lung<sup>32–34</sup> | Bronchial columnar cell dysplasia; Squamous dysplasia/carcinoma in situ; Atypical adenomatous hyperplasia; Diffuse idiopathic pulmonary neuroendocrine cell hyperplasia; HPV-related respiratory papillomatosis; Pulmonary fibrosis |
| Stomach | Gastric epithelial dysplasia; Gastric adenoma; Intestinal metaplasia; Helicobacter pylori; Chronic atrophic gastritis |
| Esophagus<sup>35</sup> | Barrett's esophagus; Achalasia; Atrophic gastritis; Esophagitis of varying types |
| Bladder<sup>36</sup> | Carcinoma in situ (also called CIS or Tis); Urothelial hyperplasia; Urothelial papilloma; Urothelial dysplasia; Intestinal metaplasia; Keratinising squamous metaplasia; Condyloma acuminatum |
| Liver<sup>37</sup> | Inflammatory liver disease; Dysplastic nodules (DN); Monoclonal gammopathy of unknown significance; Hepatitis B virus (HBV); Hepatitis C virus (HCV) |
| Kidney<sup>38</sup> | Dysplasia in the tubules adjacent to RCC; Atypical hyperplasia in the cyst epithelium of von Hippel-Lindau syndrome; Adenoma; Nephroblastomatosis |
| Pancreas<sup>39,40</sup> | Mucinous cystic neoplasm (MCN); Intraductal papillary mucinous neoplasm (IPMN); Solid pseudopapillary neoplasm (SPN); Neuroendocrine tumors; Acute and chronic pancreatitis |
| Breast<sup>41</sup> | Atypical hyperplasia; Ductal carcinoma in situ (DCIS); Benign neoplasm of breast; Lobular carcinoma in situ; Sclerosing adenosis |
| Cervical | Cervical intraepithelial neoplasia [CIN], grade I. Low grade squamous intraepithelial lesion (LSIL); CIN, grade II; CIN III; Dysplasia of cervix uteri, unspecified |
| Vaginal | Vaginal Intraepithelial Neoplasia 1 (VAIN 1); VAIN 2; VAIN 3 |
| Vulva | Vulvar Intraepithelial Neoplasia 1 (VIN 1); VIN 2; VIN 3 |
| Anal | Low-grade squamous cell intraepithelial lesions (LSILs) - anal intraepithelial neoplasia (AIN 1); HSIL - AIN 2; HSIL - AIN 3 |
| Penis<sup>42</sup> | Penile intraepithelial neoplasia (PeIN); Balanitis xerotica obliterans (BXO); Buschke-Lowenstein tumour bowenoid papulosis; Leukoplakia |

## Notes

### Competing Interest Statement

The authors have declared no competing interest.

### Author Declarations

This study was approved by the University of Florida (UF) Institutional Review Board.

